# Effect of JYNNEOS vaccination on mpox clinical progression among cisgender male cases with confirmed infection

**DOI:** 10.1101/2025.01.21.25320915

**Authors:** Lauren Granskog, Kayla Saadeh, Kieran Lorenz, Joshua Quint, Joshua Vance, Tarek Salih, Timothy Lo, Kathleen Jacobson, Marisa Ramos, Philip Peters, Eric Chapman, Robert E. Snyder, Joseph A. Lewnard

## Abstract

**Background:** The JYNNEOS modified vaccinia virus Ankara vaccine has proven effective in preventing clade IIb mpox disease during the ongoing global outbreak associated with sexual transmission. However, understanding of the effect of vaccination on mpox clinical presentation remains limited.

**Methods:** We interviewed cisgender males with confirmed mpox (cases) reported to the California Department of Public Health from May 2022 through December 2023. We ascertained cases’ vaccination statuses via the California Immunization Registry. We estimated JYNNEOS vaccine effectiveness against progression (VEP) to disease involving disseminated lesions via the adjusted odds ratio of vaccination, comparing cases who reported lesions disseminated across multiple anatomical regions to cases who reported lesions contained to a single anatomical region. We used the same approach to estimate VEP for prodromal symptoms.

**Findings:** Analyses included 4,613 cases, among whom 3,045 (66.0%) reported disseminated lesions and 1,956 (42.4%) had HIV infection. Among cases who reported disseminated lesions, 114 (3.7%) received pre-exposure vaccination and 43 (1.4%) received post-exposure vaccination only. In contrast, among cases who reported lesions contained to a single anatomical region, 286 (18.2%) received pre-exposure vaccination and 146 (9.3%) received post-exposure vaccination only. For any pre-exposure vaccination, VE*_P_* was 58.9% (95% confidence interval: 50.4-65.9%), while VE*_P_* for two pre-exposure doses was 61.0% (47.3-71.1%). For post-exposure vaccination, VE*_P_* was 15.9% (3.3-26.8%). Among cases without HIV and cases with HIV, pre-exposure VE*_P_* was 66.4% (56.6-74.0%) and 45.3% (28.0-58.5%), respectively. Pre-exposure vaccination was also associated with reduced odds for illness involving fever, chills, and lymphadenopathy.

**Interpretation:** Among cisgender males with mpox, pre-exposure vaccination with JYNNEOS is associated with less severe illness. Awareness of an attenuated disease phenotype involving localized lesions without accompanying prodromal symptoms is needed among providers to ensure accurate diagnosis of mpox in previously-vaccinated individuals.

## BACKGROUND

Since 2022, a global outbreak of clade IIb human mpox virus (hMPXV) has caused >100,000 confirmed mpox infections in regions where the disease was not previously endemic (1). This outbreak has been primarily associated with sexual transmission, with cases concentrated among gay, bisexual, and other men who have sex with men (MSM; 2,3). In 2022, the US Food and Drug Administration (FDA) issued emergency use authorization for vaccination with JYNNEOS to prevent mpox (4). JYNNEOS vaccine is a third-generation, live-attenuated vaccine derived from a modified vaccinia Ankara virus lineage, originally licensed to prevent smallpox.

National immunization technical advisory groups of multiple countries currently recommend vaccination with JYNNEOS for people at high risk of exposure to hMPXV. In the US, recommendations of the Advisory Committee on Immunization Practices encompass MSM as well as transgender or nonbinary people who were diagnosed with any sexually-transmitted infection or who had multiple sexual partners within 6 months; people who have had sex in a commercial venue or large public event within 6 months; people who anticipate having any of these exposures; and sexual partners of individuals with these risk factors (5).

Observational studies have demonstrated that JYNNEOS is effective in preventing confirmed cases of mpox during the ongoing clade IIb outbreak (6–9). Such studies have also reported lower risk of hospitalization among vaccinated cases in comparison to unvaccinated cases (10–12). However, severe disease necessitating hospitalization has occurred in a small fraction (≤6%) of mpox cases outside sub-Saharan Africa (10,13). Understanding vaccine effects on a broader spectrum of mpox clinical presentations is therefore needed, including to inform clinical awareness of the need for testing among persons experiencing less-severe manifestations who remain at risk of transmitting infection (14). We used data collected from standardized disease surveillance interviews reported to the California Department of Public Health (CDPH) to assess JYNNEOS vaccination history among persons experiencing disease involving differing clinical manifestations.

## METHODS

### Study population and data collection

Public health interviews were conducted by telephone for persons with confirmed mpox (“cases”) reported to CDPH between May 12, 2022 and December 31, 2023. Reporting is mandatory for all persons with hMPXV or orthopoxvirus DNA detected by polymerase chain reaction testing from a cutaneous lesion swab. For each case, index dates were defined as the earliest of the following time-stamped fields within surveillance records from providers, laboratories, or case interviews: symptom onset date, specimen collection date, date of specimen receipt by the laboratory, or date of receipt of the laboratory report by CDPH. We restricted analyses to cases among cisgender males due to prohibitively low sample sizes among females and gender-minority populations. Because clinical testing of specimens other than cutaneous lesion swabs is not approved, the study was implicitly restricted to cases with symptomatic illness involving ≥1 lesion.

Standardized interview forms were adapted from the US Centers for Disease Control and Prevention (CDC) case report form (15). Structured data were collected on the presence or absence of lesions at pre-specified anatomical sites (head, neck, mouth, face, arms, palms, legs, soles of feet, trunk, genitals, peri-anal area). Interviewers also had the option to record data on lesions at “other” sites within unstructured fields. Cases with lesion data reported only in the unstructured “other” field were excluded from analyses, as it was not possible to verify whether lesions were absent from anatomical sites not mentioned in this field without accompanying structured data entries. Cases were also asked to report whether they experienced fever, chills, lymphadenopathy, and pruritis during their illness.

Further demographic and behavioral variables included sex, age, race/ethnicity, sexual orientation and gender identity, gender of sex partners, and sexual contact within the three weeks preceding cases’ index date. Data collected from interviews undertaken by local health jurisdictions were submitted to CDPH. Cases’ HIV infection status and most recent viral load relative to their mpox episode date (for cases with HIV) were verified by linkage to CDPH Office of AIDS registry data. Receipt of JYNNEOS was verified by linkage to the California Immunization Registry, where all JYNNEOS administrations in the state of California are reported.

These analyses were considered non-human subjects research by the Committee for the Protection of Human Subjects of the California Health and Human Services Agency. Informed consent was not required for participation in public health case interviews.

### Outcomes

The primary study outcome was occurrence of disseminated lesions, defined as presence of lesions at sites across multiple anatomical regions by the time of the case interview. We grouped sites at which cases reported lesions into 6 broader anatomical regions via a modified “Rule of Nines” framework aligned with potential hMPXV exposure sites (16); these included the head, face or mouth; neck, back and torso; arms, wrists, and hands/palms; legs and feet/soles; genitals; and peri-anal area and buttocks (**Figure 1**). Secondary outcomes included the number of anatomical regions at which cases reported lesions (measured continuously from 1-6) and the occurrence of fever, chills, swollen lymph nodes, and pruritis at any time during illness.

**Figure 1:**
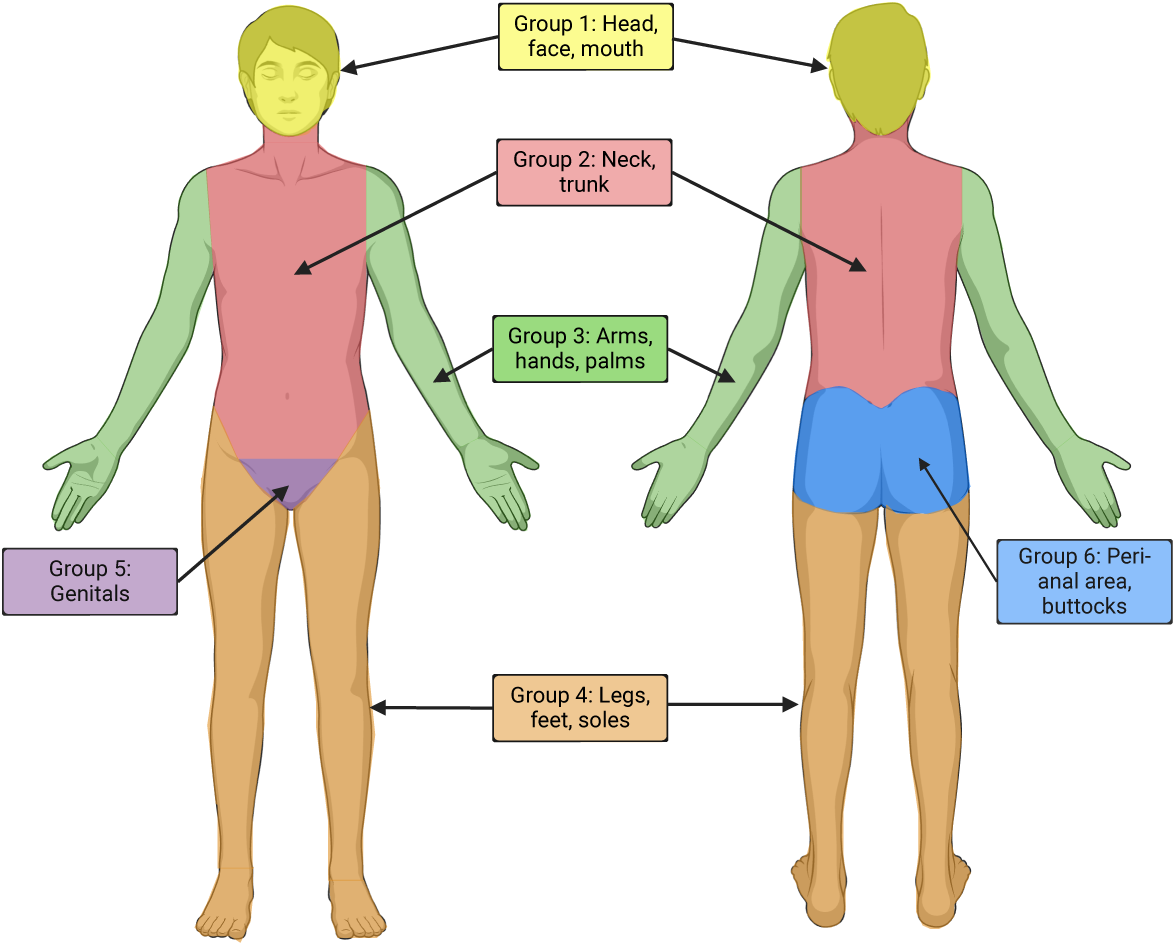
Regional groupings of anatomical sites. We delineate anatomical regions of adjacent sites at which cases reported lesions were present or not present, aiming to accommodate differences in the specificity with which cases reported specific lesion sites during interviews. Groupings were defined using a modified “Rule of Nines” framework aligned with potential exposure sites.

### Exposures

We defined pre-exposure vaccination with JYNNEOS as receipt of JYNNEOS ≥14 days before the index date. We defined pre-exposure immunization status as “fully vaccinated” if a second dose was received ≥24 days after the first dose and ≥14 days before cases’ index date (5); otherwise, we considered cases who had received one dose of JYNNEOS ≥14 days before the index date to be “partially vaccinated”. For cases without any pre-exposure JYNNEOS administration, we defined JYNNEOS administrations occurring <14 days before the index date as post-exposure vaccination. Cases with no record of receiving JYNNEOS were considered unvaccinated.

### Statistical analysis

The primary study objective was to estimate vaccine effectiveness against progression (VE*_P_*) to disease involving disseminated lesions associated with pre-exposure JYNNEOS vaccination. We measured VE*_P_* via the adjusted odds ratio (aOR) of pre-exposure vaccination with JYNNEOS among cases who reported disseminated lesions versus cases with lesions contained to a single anatomical region, defining VE_!_ = (1 − aOR) × 100%. As secondary objectives, we also computed VE*_P_* separately for cases who were either fully or partially vaccinated prior to exposure, and for cases who received JYNNEOS post-exposure; we note, however, that a lack of data on time from hMPXV exposure to JYNNEOS administration may limit the generalizability of VE*_P_* estimates for doses received post-exposure (17). We used the same framework to compute VE*_P_* against other symptoms including fever, chills, swollen lymph nodes, and pruritis as a secondary objective.

We expected that concurrent HIV infection could confound VE*_P_* estimation, as HIV infection could be associated with both vaccine access and individuals’ risk of severe illness, in addition to modifying VE*_P_* (18,19). To mitigate resulting bias, we computed the aOR of JYNNEOS receipt via conditional logistic regression models with strata defined for HIV infection status. Restricting analyses to confirmed cases was expected to mitigate other confounding pathways potentially driven by associations of vaccination and mpox outcomes with characteristics predicting exposure such as age, race/ethnicity, and sexual orientation, or history of sexual contact with male partners (**Figure S1**). Tecovirimat receipt was not considered a confounder due to the restriction of tecovirimat eligibility to patients already experiencing the primary study outcome (disseminated lesions affecting ≥25% of the body surface) under the CDC Expanded Access Investigational New Drug Protocol (20).

We also aimed to determine whether HIV infection modified the effectiveness of JYNNEOS against disease progression. We repeated analyses within subgroups including HIV-negative cases and cases with HIV, and within subgroups of cases with HIV whose most recent CD4 counts (reported to CDPH within 12 months before the index date) fell within the ranges of ≥500 or <500 per mm^3^. Cases without any CD4 count reported to CDPH within this timeframe were considered to have an unknown CD4 count.

To better understand the relationship between vaccination and lesion extent, we also computed aORs relating vaccination with JYNNEOS to the number of anatomical regions at which cases reported lesions on a continuous scale. Similar to the analyses described above, we used conditional logistic regression models defining strata for HIV infection to mitigate confounding.

Last, as an exploratory analysis aiming to understand HIV as an independent risk factor for disease progression, we estimated the aOR of HIV infection among cases who reported disseminated lesions versus cases with lesions contained to a single anatomical region. We compared cases with HIV to cases without HIV, and also compared cases with HIV with CD4 counts ≥500/mm^3^, <500/mm^3^, or unknown CD4 counts to cases without HIV. We computed aORs via conditional logistic regression models with strata defined for receipt of JYNNEOS (pre-exposure or post-exposure).

## RESULTS

In total, 6,166 cases with confirmed mpox were reported to CDPH between May 12, 2022 and December 31, 2023, among whom 5,767 (94.3% of 6,116) were cisgender males (**Figure 2**). The analytic population included 4,613 cisgender male cases (80.0% of 5,767) for whom comprehensive answers to questions about lesion dissemination were recorded from public health surveillance investigations. Among these cases, 2,056 (44.6%) identified as Hispanic or Latino, 1,397 (30.2%) as non-Hispanic White, 545 (11.8%) as non-Hispanic Black, and 264 (5.7%) as non-Hispanic Asian (**Table 1**), closely matching demographics of cases reported statewide (**Table S1**). The greatest share of cases (1,845; 40.0%) were 26-35 years old, 3,817 (82.7%) identified as gay or same-gender loving, and 3,319 (71.9%) disclosed sexual contact with at least ≥1 partner within three weeks before their illness. Altogether, 1,956 (42.4%) cases were living with HIV, among whom 935 had CD4 counts ≥500/mm^3^, 671 had CD4 counts <500/mm^3^, and 350 had unknown CD4 counts (47.8%, 34.3%, and 17.9% of 1,956, respectively). In total, 832 (18.0%) cases were documented to have received tecovirimat (**Table S2**).

**Figure 2:**
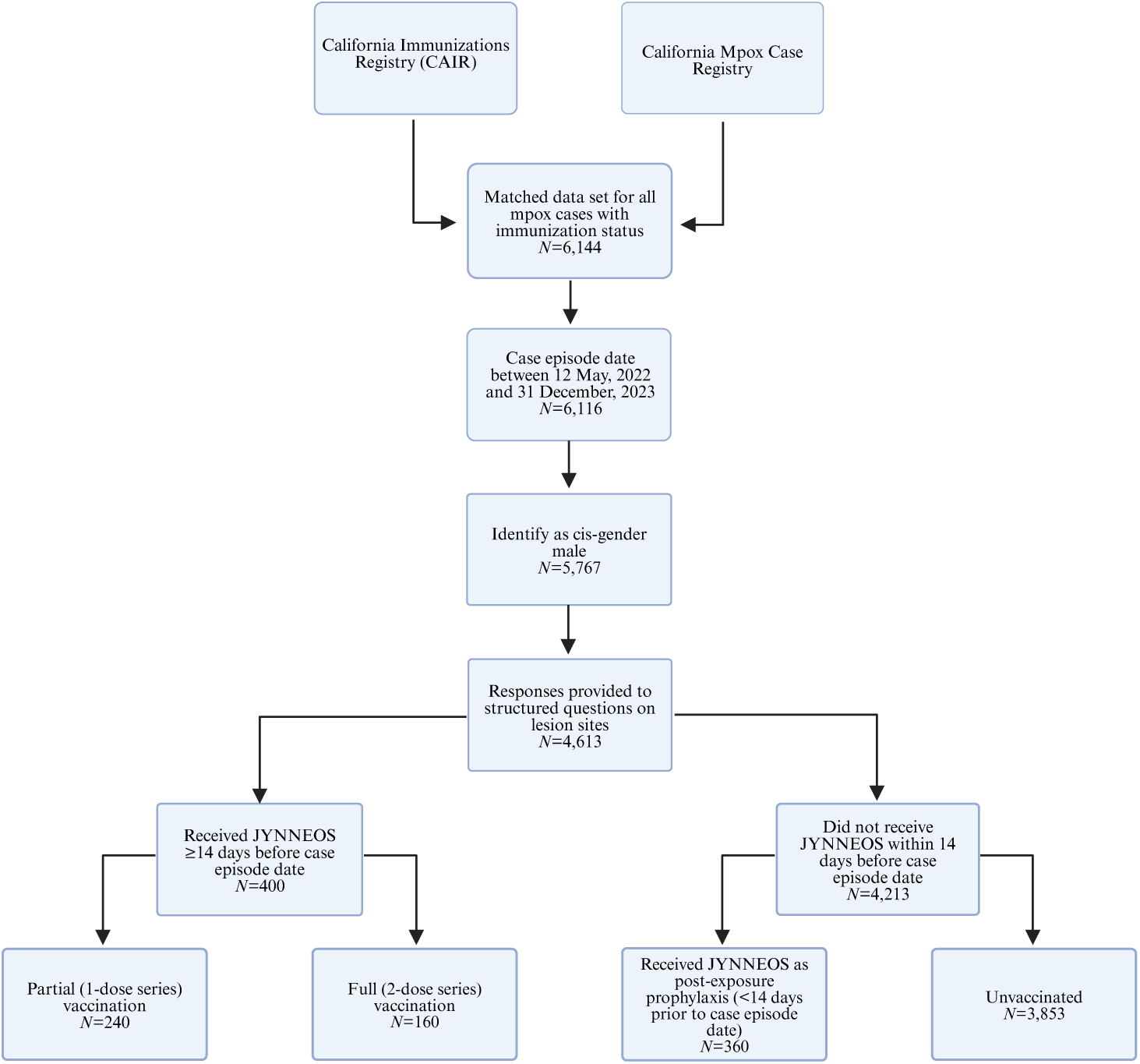
Study flowchart. We detail the number of cases retained after application of exclusion criteria based on case episode date, sex/gender, and availability of data on the primary study outcome, as well as the number of subjects belonging to each classification for JYNNEOS vaccine exposure.

**Table 1:** Characteristics of the study population.

| Characteristic |  | Participants, by history of JYNNEOS vaccination, n (%) |  |  |  |  |
| --- | --- | --- | --- | --- | --- | --- |
|  |  | Any vaccination history | Full (2 dose) pre-exposure series<sup>1</sup> | Partial (1 dose) pre-exposure series<sup>1</sup> | Post-exposure vaccination only<sup>2</sup> | Unvaccinated |
|  |  | N=4,613 | N=160 | N=240 | N=360 | N=3,853 |
| Age (years) | ≤15 | 13 (0.3) | 0 | 0 | 0 | 13 (0.3) |
|  | 16-25 | 461 (10.0) | 12 (7.5) | 21 (8.8) | 23 (6.4) | 405 (10.5) |
|  | 26-35 | 1,845 (40.0) | 60 (37.5) | 105 (43.8) | 122 (33.9) | 1558 (40.4) |
|  | 36-45 | 1,368 (29.7) | 52 (32.5) | 63 (26.2) | 124 (34.4) | 1129 (29.3) |
|  | 46-55 | 644 (14.0) | 22 (13.8) | 33 (13.8) | 64 (17.8) | 525 (13.6) |
|  | 56-65 | 256 (5.5) | 13 (8.1) | 17 (7.1) | 24 (6.7) | 202 (5.2) |
|  | ≥65 | 26 (0.6) | 1 (0.6) | 1 (0.4) | 3 (0.8) | 21 (0.5) |
| Race/ethnicity | Hispanic or Latino | 2,056 (44.6) | 56 (35.0) | 87 (36.2) | 136 (37.8) | 1777 (46.1) |
|  | White, non-Hispanic | 1,397 (30.3) | 75 (46.9) | 109 (45.4) | 143 (39.7) | 1070 (27.8) |
|  | Black or African American, non-Hispanic | 545 (11.8) | 6 (3.8) | 12 (5.0) | 23 (6.4) | 504 (13.1) |
|  | Asian, non-Hispanic | 264 (5.7) | 7 (4.4) | 20 (8.3) | 24 (6.7) | 213 (5.5) |
|  | Native Hawaiian/other Pacific Islander, non-Hispanic | 22 (0.5) | 0 | 2 (0.8) | 3 (0.8) | 17 (0.4) |
|  | American Indian or Alaska Native, non-Hispanic | 21 (0.5) | 0 | 0 | 2 (0.6) | 19 (0.5) |
|  | Multiple or other races or ethnicities | 147 (3.2) | 9 (5.6) | 6 (2.5) | 17 (4.7) | 115 (3.0) |
|  | Unknown | 161 (3.5) | 7 (4.4) | 4 (1.7) | 12 (3.3) | 138 (3.6) |
| HIV infection status | Living with HIV infection | 1,956 (42.4) | 44 (27.5) | 85 (35.4) | 161 (44.7) | 1666 (43.2) |
|  | HIV negative | 2,657 (57.6) | 116 (72.5) | 155 (64.6) | 199 (55.3) | 2187 (56.8) |
| Male-male sexual contact within 3 weeks preceding episode | Yes | 3,319 (71.9) | 144 (90.0) | 203 (84.6) | 300 (83.3) | 2672 (69.3) |
|  | No | 1,025 (22.2) | 10 (6.2) | 24 (10.0) | 47 (13.1) | 944 (24.5) |
|  | Unknown | 269 (5.8) | 6 (3.8) | 13 (5.4) | 13 (3.6) | 237 (6.2) |
| Sexual orientation | Heterosexual/straight | 323 (7.0) | 2 (1.2) | 5 (2.1) | 5 (1.4) | 311 (8.1) |
|  | Gay, lesbian, or same-gender loving | 3,343 (72.5) | 140 (87.5) | 206 (85.8) | 309 (85.8) | 2688 (69.8) |
|  | Bisexual | 474 (10.3) | 8 (5.0) | 10 (4.2) | 17 (4.7) | 439 (11.4) |
|  | Questioning | 4 (0.1) | 0 | 0 | 1 (0.3) | 3 (0.1) |
|  | Declined to answer | 113 (2.4) | 2 (1.2) | 6 (2.5) | 3 (0.8) | 102 (2.6) |
|  | Unknown | 356 (7.7) | 8 (5.0) | 13 (5.4) | 25 (6.9) | 310 (8.0) |
| Tecovirimat receipt | Tecovirimat received | 832 (18.0) | 25 (15.6) | 40 (16.7) | 76 (21.1) | 691 (17.9) |
|  | Tecovirimat not received | 3,781 (82.0) | 135 (84.4) | 200 (83.3) | 284 (78.9) | 3,162 (82.1) |

<sup>1</sup>Individuals were considered to have received pre-exposure vaccination series if at least one dose was received ≥14 days before the case episode date. Individuals were considered to have received a full series if a first pre-exposure dose was received ≥24 days before the second pre-exposure dose.
<sup>2</sup>Individuals who received any dose within 14 days of the case episode date were considered to have received vaccination as post-exposure prophylaxis.

Disseminated lesions were reported by 3,045 cases (66.0% of 4,613 cases), including 1,627 HIV-negative cases (61.2% of 2,657 HIV-negative cases) and 1,418 cases with HIV (72.5% of 1,956 cases with HIV; **Figure 3**). Among cases with HIV, disseminated lesions were reported by 71.1% of cases with CD4 counts ≥500/mm^3^ (665/935) and 73.5% of cases with CD4 counts <500/mm^3^ (493/671), as well as 74.3% of cases with unknown CD4 counts (260/350). Overall, 2,013 (43.6%) cases reported lesions on their face, mouth, or head; 1,789 (38.8%) on their neck or trunk; 2,279 (49.4%) on their hands or arms; 2,525 (54.7%) on their genitals; 1,481 (32.1%) near their anus or on their buttocks; and 1,871 (40.6%) on their legs or feet (**Table 2**). Most cases who received tecovirimat reported experiencing disseminated lesions (590/832, 70.9%), although receipt of tecovirimat was not strongly associated with this outcome after adjustment for JYNNEOS vaccination and HIV infection status (aOR=1.08 [0.98-1.20]; **Table S2**).

**Figure 3:**
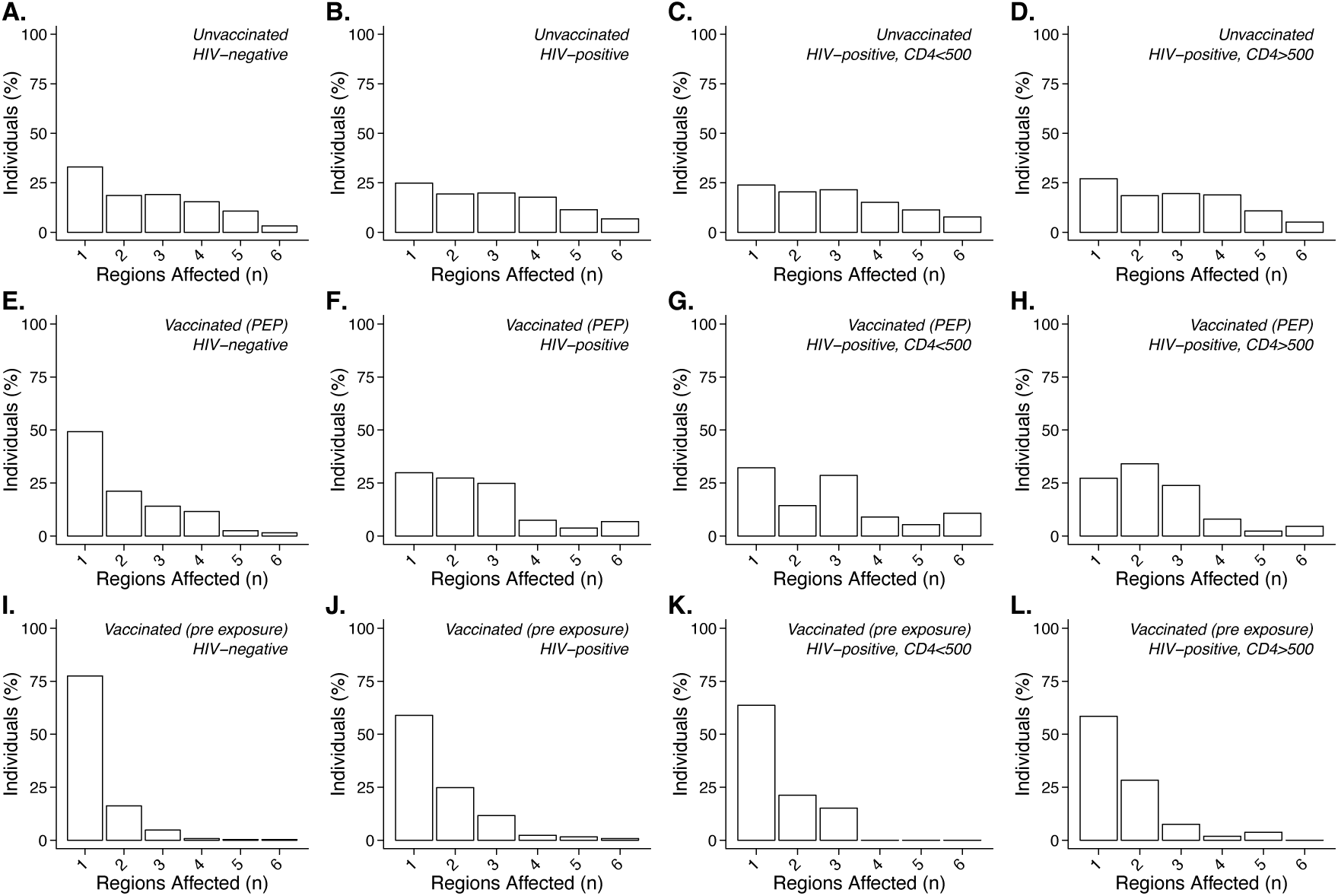
Distribution of the number of anatomical regions affected by lesions, according to vaccination history and HIV infection status. We plot the proportion of cases reporting lesions contained to a single anatomical region or disseminated across 2, 3, 4, 5, or 6 anatomical regions, stratified by cases’ history of vaccination (unvaccinated, panels **A**-**D**; vaccinated as post-exposure prophylaxis only, panels **E**-**H**; or vaccinated with 1 or 2 pre-exposure doses, panels **I**-**L**) and HIV infection status (from left to right, HIV-negative cases, cases living with HIV, cases living with HIV with CD4 counts <500/mm^3^, and cases living with HIV with CD4 counts ≥500/mm^3^).

**Table 2:**
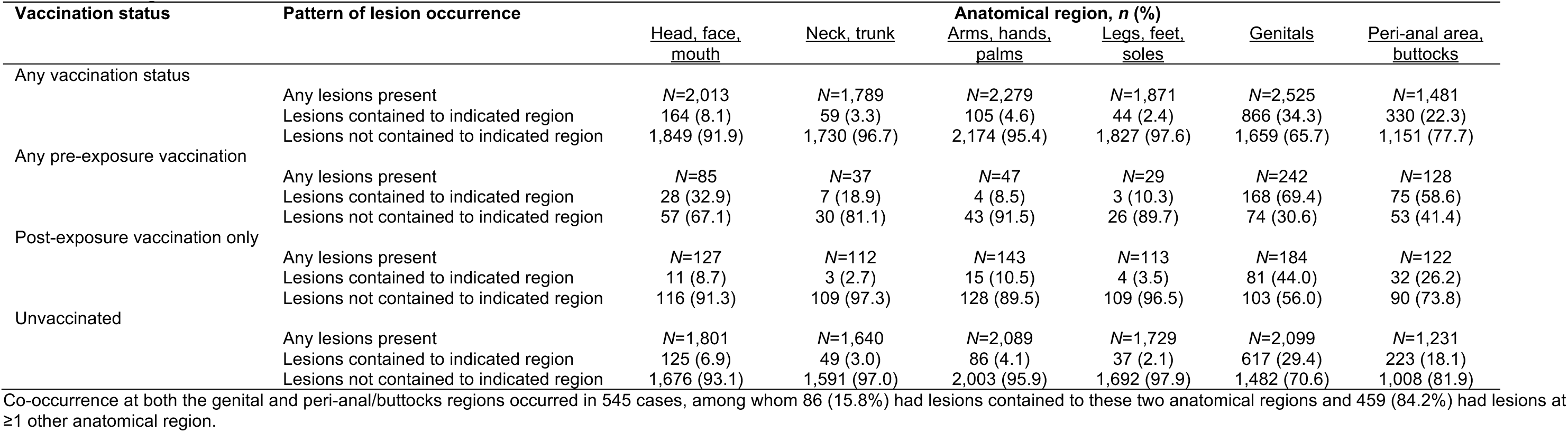
Lesion locations among cases experiencing lesions contained to single anatomical regions or disseminated lesions across multiple anatomical regions.

Overall, 114 of 3,045 cases who reported disseminated lesions (3.7%) received pre-exposure vaccination with JYNNEOS, as compared to 286 of 1,568 cases with lesions contained to a single anatomical region (18.2%; **Table 3**). For any pre-exposure vaccination with JYNNEOS, VE*_P_* was 58.9% (95% confidence interval: 50.4-65.9%). For partial and full pre-exposure vaccination series, VE*_P_* was 57.6% (46.3-66.5%) and 61.0% (47.3-71.1%), respectively. Among cases who did not receive pre-exposure JYNNEOS vaccination, 43 cases reporting disseminated lesions (1.5% of 2,931) and 146 cases with lesions contained to a single anatomical region (11.4% of 1,282) received JYNNEOS as post-exposure vaccination, for which VE*_P_* was 15.9% (3.3-26.8%). With each additional anatomical region at which cases reported lesions, cases had 57.2% (51.7-62.2%) lower adjusted odds of having received pre-exposure vaccination with JYNNEOS and 20.8% (14.5-26.6%) lower adjusted odds of having received JYNNEOS post-exposure (**Table S3**).

**Table 3:** JYNNEOS vaccine effectiveness against progression (VE*_P_*) to disseminated lesion occurrence.

| Vaccination status<br>*P* | Pattern of lesion occurrence |  | VE<sub>P</sub>, % (95% CI) |
| --- | --- | --- | --- |
|  | Disseminated lesions<br>across multiple<br>anatomical regions, *n* (%) | Lesions contained to a<br>single anatomical<br>region, *n* (%) |  |
|  | *N*=3,045 | *N*=1,568 |  |
| Unvaccinated | 2,717 (89.2) | 1,136 (72.4) | ref. |
| Any pre-exposure immunization | 114 (3.7) | 286 (18.2) | 58.9 (50.4, 65.9) |
| Partial series | 71 (2.3) | 169 (10.8) | 57.6 (46.3, 66.5) |
| Full series | 43 (1.4) | 117 (7.5) | 61.0 (47.3, 71.1) |
| Immunization as post-exposure prophylaxis alone | 214 (7.0) | 146 (9.3) | 15.9 (3.3, 26.8) |

CI: Confidence interval; VE<sub>P</sub>: Vaccine effectiveness against progression to disseminated lesions. We define VE<sub>P</sub> as (1–adjusted odds ratio)×100%, for the adjusted odds ratio of each vaccine exposure versus no vaccination comparing cases who experienced disseminated lesions to those with lesions contained to a single region. We estimate the adjusted odds ratio via conditional logistic regression, matching participants on HIV infection status.

For pre-exposure JYNNEOS series, VE*_P_* was 66.4% (56.6-74.0%) among cases without HIV and 45.3% (28.0-58.5%) among cases with HIV (**Table 4**). For JYNNEOS delivered post-exposure, VE*_P_* was 24.2% (7.3-38.1%) among cases without HIV and 6.6% (–13.2-23.0%) among cases with HIV. Stratified analyses did not identify appreciable differences in VE*_P_* among cases with CD4 counts ≥500/mm^3^ and <500/mm^3^ for any pre-exposure series (VE*_P_*=42.9% [13.1-63.0%] and 52.2% [14.1-73.0%], respectively). For cases with unknown CD4 count, VE*_P_* was 44.2% (10.1-65.5%) for any pre-exposure series. Post-exposure receipt of JYNNEOS was not strongly associated with protection from progression within any CD4 count-based stratum among cases with HIV.

**Table 4:** JYNNEOS vaccine effectiveness against progression (VE*_P_*) to disseminated lesion occurrence, according to cases’ HIV infection status.

| HIV infection status | Vaccination status | Pattern of lesion occurrence |  | VE<sub>P</sub>, % (95% CI) |
| --- | --- | --- | --- | --- |
|  |  | Disseminated lesions across multiple anatomical regions, *n* (%) | Lesions contained to a single anatomical region, *n* (%) |  |
| HIV-negative |  | *N*=1,627 | *N*=1,030 |  |
|  | Unvaccinated | 1,465 (90.0) | 722 (70.1) | ref. |
|  | Any pre-exposure immunization | 61 (3.7) | 210 (20.4) | 66.4 (56.6, 74.0) |
|  | Partial (1-dose) pre-exposure series | 37 (2.3) | 118 (11.5) | 64.4 (50.6, 74.3) |
|  | Full (2-dose) pre-exposure series | 24 (1.5) | 92 (8.9) | 69.1 (53.8, 79.4) |
|  | Immunization as post-exposure prophylaxis alone | 101 (6.2) | 98 (9.5) | 24.2 (7.3, 38.1) |
| Living with HIV infection: all |  | *N*=1,418 | *N*=538 |  |
|  | Unvaccinated | 1,252 (88.3) | 414 (77.0) | ref. |
|  | Any pre-exposure immunization | 53 (3.7) | 76 (14.1) | 45.3 (28.0, 58.5) |
|  | Partial (1-dose) pre-exposure series | 34 (2.4) | 51 (9.5) | 46.8 (25.2, 62.1) |
|  | Full (2-dose) pre-exposure series | 19 (1.3) | 25 (4.6) | 42.5 (9.6, 63.5) |
|  | Immunization as post-exposure prophylaxis alone | 113 (8.0) | 48 (8.9) | 6.6 (−13.2, 23.0) |
| Living with HIV infection: CD4 count ≥500/mm<sup>3</sup> |  | *N*=665 | *N*=270 |  |
|  | Unvaccinated | 579 (87.1) | 215 (79.6) | ref. |
|  | Any pre-exposure immunization | 22 (3.3) | 31 (11.5) | 42.9 (13.1, 63.0) |
|  | Partial (1-dose) pre-exposure series | 16 (2.4) | 27 (10.0) | 49.1 (15.1, 69.3) |
|  | Full (2-dose) pre-exposure series | 6 (0.9) | 4 (1.5) | 17.7 (−83.0, 63.3) |
|  | Immunization as post-exposure prophylaxis alone | 64 (9.6) | 24 (8.9) | 0.3 (−28.3, 23.2) |
| Living with HIV infection: CD4 count <500/mm<sup>3</sup> |  | *N*=493 | *N*=178 |  |
|  | Unvaccinated | 443 (89.9) | 139 (78.1) | ref. |
|  | Any pre-exposure immunization | 12 (2.4) | 21 (11.8) | 52.2 (14.1, 73.0) |
|  | Partial (1-dose) pre-exposure series | 12 (2.4) | 16 (9.0) | 43.7 (−0.5, 68.3) |
|  | Full (2-dose) pre-exposure series | 0 | 5 (2.8) | — |
|  | Immunization as post-exposure prophylaxis alone | 38 (7.7) | 18 (10.1) | 11.0 (−23.9, 36.0) |
| Living with HIV infection: Unknown CD4 count |  | *N*=260 | *N*=90 |  |
|  | Unvaccinated | 230 (88.5) | 60 (66.7) | ref. |
|  | Any pre-exposure immunization | 19 (7.3) | 24 (26.7) | 44.2 (10.1, 65.5) |
|  | Partial (1-dose) pre-exposure series | 6 (2.3) | 8 (8.9) | 45.8 (−21.9, 76.3) |
|  | Full (2-dose) pre-exposure series | 13 (5.0) | 16 (17.8) | 43.3 (1.0, 68.0) |
|  | Immunization as post-exposure prophylaxis alone | 11 (4.2) | 6 (6.7) | 18.4 (−50.9, 55.2) |

CI: Confidence interval; VE<sub>P</sub>: Vaccine effectiveness against progression to disseminated lesions. We define VE<sub>P</sub> as (1−adjusted odds ratio)×100%, for the adjusted odds ratio of each vaccine exposure versus no vaccination comparing cases who experienced disseminated lesions to those with lesions contained to a single region. We estimate the adjusted odds ratio via conditional logistic regression, matching participants on HIV infection status.

Among all 4,613 cases included in analyses, 4,490 (97.3%) provided responses about the occurrence of fever, 4,401 (95.3%) about chills, 4,307 (93.4%) about lymphadenopathy, and 4,124 (89.4%) about pruritis during their illness (**Table 5**). Few of the cases who reported these prodromal symptoms had received any pre-exposure JYNNEOS doses; cases who had received pre-exposure JYNNEOS doses comprised 4.3% (115/2,685) of those reporting fever, 4.0% (87/2,150) of those reporting chills, 7.1% (150/2,118) of those reporting lymphadenopathy, and 9.3% (209/2,245) of reporting pruritis. Receipt of pre-exposure JYNNEOS doses was more common among cases who reported that they did not experience prodromal symptoms. Cases who had received pre-exposure JYNNEOS doses comprised 15.1% (272/1,805), 13.3% (300/2,251), 10.4% (228/2,189), and 9.2% (173/1,879) of those who reported that they did not experience fever, chills, lymphadenopathy, and pruritis, respectively. For pre-exposure JYNNEOS series, VE*_P_* against illness involving fever, chills, lymphadenopathy, and pruritis was 52.8% (43.1-60.9%), 57.1% (46.7-65.4%), 22.1% (8.0-34.1%), and –0.6% (–16.1-12.8%), respectively. Findings were similar in analyses stratified by cases’ HIV infection status (**Table S4**).

**Table 5:**
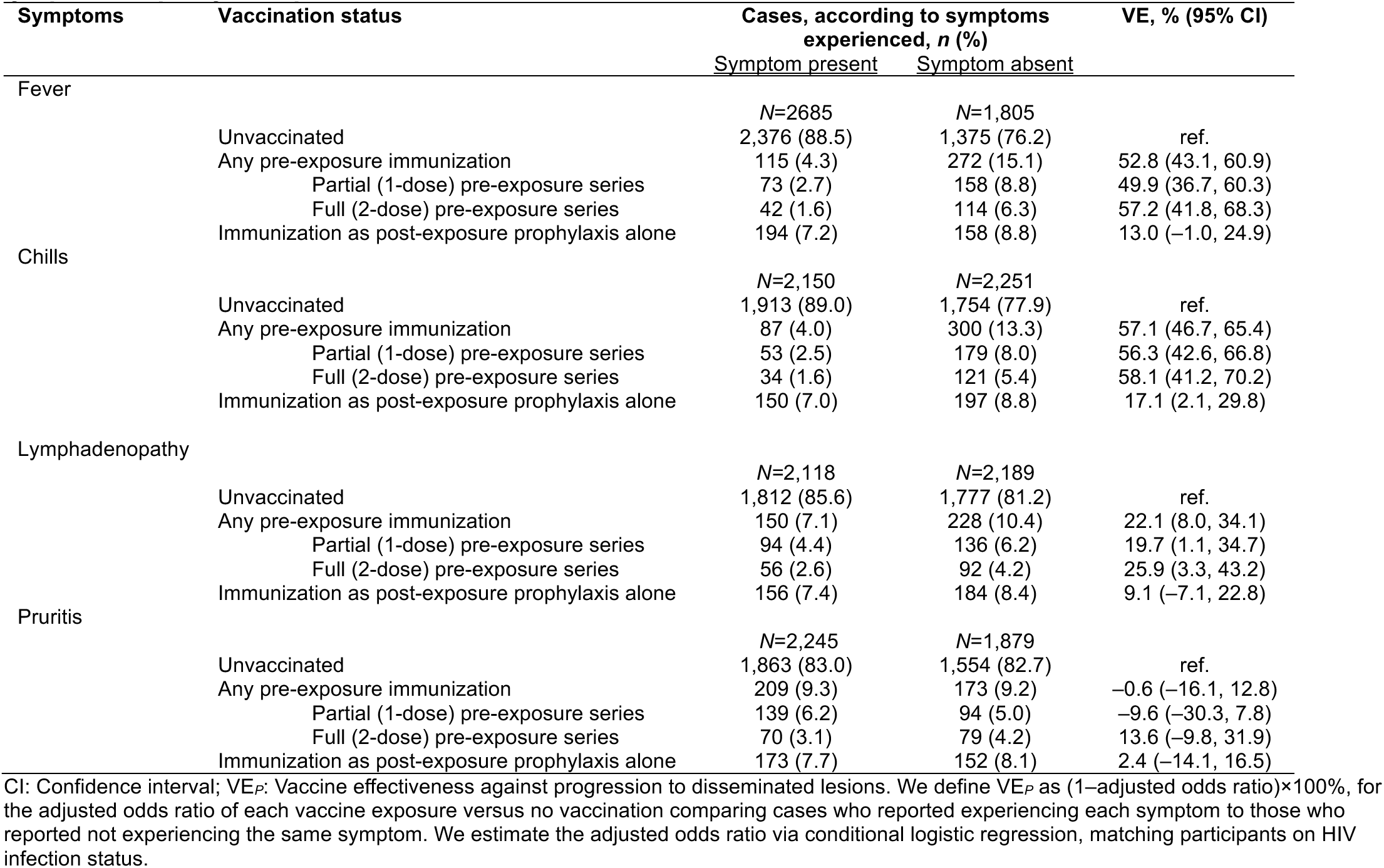
JYNNEOS vaccine effectiveness against progression (VE*_P_*) to illness involving fever, chills, lymphadenopathy, and pruritis.

In exploratory analyses, HIV infection was associated with increased risk of progression to disseminated lesions regardless of vaccination status. Compared to cases with lesions contained to a single anatomical region, cases who reported disseminated lesions had 1.16-fold (1.08-1.24) higher adjusted odds of HIV infection (**Table 6**). This association was apparent within each stratum based on JYNNEOS vaccine receipt; among unvaccinated cases, cases who reported disseminated lesions had 1.12 (1.04-1.21) fold higher adjusted odds of HIV infection, while among recipients of 1 and 2 pre-exposure doses, adjusted odds of HIV infection were 1.68 (1.05-2.67) and 2.09 (1.14-3.81) fold higher, respectively, among cases who reported disseminated lesions. Among cases who received JYNNEOS as post-exposure vaccination, cases reporting disseminated lesions had 1.38 (1.06-1.81) fold higher adjusted odds of HIV infection. However, reporting fever, chills, lymphadenopathy, or pruritis was not strongly associated with cases’ HIV infection status (**Table S5**).

## DISCUSSION

Among cisgender males with laboratory-confirmed mpox, pre-exposure vaccination with JYNNEOS was associated with protection against progression to disease involving disseminated lesions. This effect was apparent among cases regardless of their HIV infection status. Point estimates of VE*_P_* did not differ appreciably among cases who had received either partial or full pre-exposure JYNNEOS series (58% and 61%). Pre-exposure JYNNEOS vaccination was also associated with protection against non-cutaneous symptoms including fever, chills, and lymphadenopathy. When compared to unvaccinated cases, cases who received any pre-exposure JYNNEOS doses had 22-57% lower odds of reporting these symptoms. Consistent with these results, a recent multijurisdictional study of US mpox cases also reported that mpox cases who received ≥2 JYNNEOS doses before exposure had lower likelihood of systemic illness and a lower median number of anatomical regions with rash (21). These findings demonstrate that in addition to preventing mpox (6–8), JYNNEOS may reduce disease severity. Clinical awareness that JYNNEOS alters the clinical presentation of mpox is needed to ensure that testing is made promptly available to vaccinated individuals, who may experience lesions that remain localized to a single anatomical region and present without common prodromal symptoms.

We obtained lower estimates of protection against disseminated disease for JYNNEOS when administered post-exposure. Among cases with HIV, we did not identify strong evidence that post-exposure vaccination with JYNNEOS attenuated clinical severity of mpox, although previous analyses demonstrate that timely post-exposure vaccination may remain important for preventing infection among persons reporting suspected or confirmed mpox exposure (22). Prior analyses have revealed that estimates of the effectiveness of post-exposure JYNNEOS vaccination may encounter bias if administration occurs only before confirmation of infection, as cases’ perceived clinical status may influence providers’ likelihood of recommending administering post-exposure vaccine doses (17,23). While restricting our sample to confirmed cases may partially address this problem, analogous challenges may persist if JYNNEOS is preferentially administered to individuals not yet experiencing severe disease manifestations, or alternatively to those who are already experiencing severe illness. Thus, our analyses may over-or under-estimate VE*_P_* for post-exposure JYNNEOS vaccination.

Within our study population, cases with HIV infection had higher odds of reporting disseminated lesions, regardless of vaccination status. Previous studies have implicated HIV infection as an important risk factor for severe disease outcomes including hospitalization and mortality (13,24), but have had insufficient sample sizes to demonstrate JYNNEOS is effective among people with HIV (6,25,26). Thus, our finding that pre-exposure JYNNEOS vaccination is protective against disease progression among cases with HIV is reassuring, and supports existing recommendations for vaccination of all individuals at risk for hMPXV exposure (5). Few cases in our study population met clinical definitions for AIDS (CD4 count <200/mm^3^), potentially restricting our ability to investigate CD4 count as a modifier of VE*_P_*.

Our analysis has several limitations. First, a substantial proportion of hMPXV infections may be asymptomatic, even among unvaccinated individuals (27,28). Because testing is currently restricted to cutaneous lesion swabs, all cases in our study were symptomatic, and our analyses may not account fully for differences in clinical outcomes of hMPXV infection among vaccinated and unvaccinated persons. Second, while it is possible that cases could go on to experience solicited symptoms at later points in time after their interview, it is unlikely that this happened for an appreciable number of cases. Median time from symptom onset to testing among US cases has been 11 days (29), median test turnaround times exceed 1 day (30), and cases are approached for public health interviews only after their diagnoses are reported to local health jurisdictions. Thus, most cases were likely interviewed during later stages of illness or during convalescence. Third, unmeasured confounding may be present under our observational study design. While restricting analyses to diagnosed cases likely mitigates bias that would result from differential healthcare-seeking among individuals who did or did not receive JYNNEOS (**Figure S1**), it remains possible that thresholds for care-seeking differ among vaccinated and unvaccinated individuals, contributing to the detection of less-severe infections among vaccinated cases. Fourth, defining the outcome of disseminated lesions may pose challenges, as presence of lesions at multiple anatomical sites may result from primary exposure occurring at multiple sites or from clinical progression (31). However, containment of lesions to body regions not associated with insertive or receptive sex acts was rare among both vaccinated and unvaccinated persons (**Table 2**). This suggests presentations of lesions at such locations reflected true rash dissemination. Fifth, JYNNEOS administrations occurring outside the state of California could not be verified from the California Immunization Registry. Sixth, our study did not address routes of vaccine administration; previous analyses have indicated that vaccinated cases experienced less-severe illness regardless of whether doses were administered intradermally or subcutaneously (21). Last, as clade I hMPXV was not detected in the United States during the study period, our study population is likely restricted to mpox cases infected with clade II hMPXV. Measuring effects of JYNNEOS on risk of infection and symptoms of clade I hMPXV remains an important priority due to recent evidence of its spread in MSM sexual networks in the Democratic Republic of the Congo (32), as well as its introduction into other settings including California.

While attenuation of mpox clinical severity by JYNNEOS offers the advantage of mitigating disease burden, the occurrence of milder disease among vaccinated cases may pose challenges to case ascertainment and interruption of transmission. Contact with lesions is not necessary for hMPXV transmission to occur: pre-symptomatic transmission has been documented (31,33) and hMPXV can be cultured from superficial samples during pre-symptomatic stages of infection (34) and during asymptomatic infection (28). Because vaccination is associated not only with a reduced extent of lesion spread, but also with reduced likelihood for cases to experience prodromal symptoms such as fever, chills, and lymphadenopathy, timely diagnosis could be more challenging among vaccinated cases, potentially impacting their ability to take precautionary measures to avoid onward transmission. Reduced clinical severity among vaccinated cases may account for the low proportion of mpox cases who reported being aware of any prior exposure to hMPXV in a recent study undertaken after JYNNEOS rollout (31).

In conclusion, our study identifies attenuation of mpox clinical severity among male recipients of any pre-exposure JYNNEOS vaccination who were subsequently diagnosed with mpox. This effect of vaccination on disease progression was apparent among all cases, regardless of HIV infection status. Complementing previous evidence that JYNNEOS vaccination is effective in preventing hMPXV infection (6–8), our findings support existing recommendations that MSM and other persons at high risk of mpox exposure should receive two doses of JYNNEOS (5). Securing high uptake of JYNNEOS remains an important public health objective to mitigate hMPXV transmission and to lower individuals’ risk of severe disease if they are exposed. In conjunction with ensuring JYNNEOS remains accessible to prevent and mitigate the severity of mpox, updated guidance is needed to ensure healthcare providers and at-risk communities are aware of the range of clinical mpox presentations, including among vaccinated persons, to avoid missed opportunities for testing.

## Data Availability

All data produced in the present study are available upon reasonable request to the California Department of Public Health.

## ACKNOWLEDGMENTS

The study was supported by the US Centers for Disease Control & Prevention cooperative agreement PS19-1901 to the CDPH Sexually Transmitted Disease Control Branch. The findings and conclusions in this report are those of the authors and do not necessarily represent the views or opinions of the CDPH or the California Health and Human Services Agency.

---

**Figure S1:**
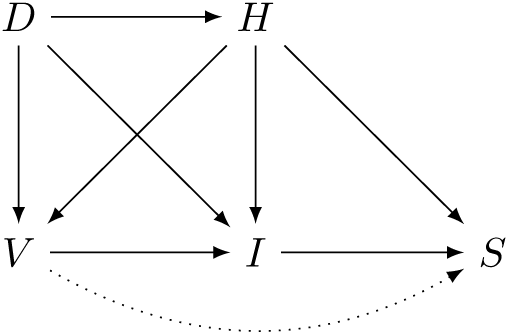
Directed acyclic graph for the confounding pathways affecting measurement of the relationship between JYNNEOS vaccination and risk of disease progression. We consider the direct effect of vaccination (*V*) on risk of progression to severe disease (*S*), VE*_P_*, which for our primary analyses is defined by presence of lesions at multiple anatomical regions. The causal direct effect of interest is illustrated via the dotted line *V*®*S.* Demographic and behavioral factors (*D*) influencing exposure to mpox, such as sexual orientation, age, gender, race, and sexual behavior preferences, are anticipated to predict individuals’ likelihood of vaccination, of infection with hMPXV (*I*), and of living with HIV infection (H). Any vaccine direct effect on susceptibility to infection is likewise expected to impact risk of hMPXV (VE*_S_* per (1)). In turn, HIV infection status, vaccination, and infection with hMPXV determine risk of severe mpox disease. This scenario implies the outcome *S* is conditionally independent of *D* given *H*, for which we adjust, and *I*, for which our case-only design selects on the condition *I=*1.

**Table S1:**
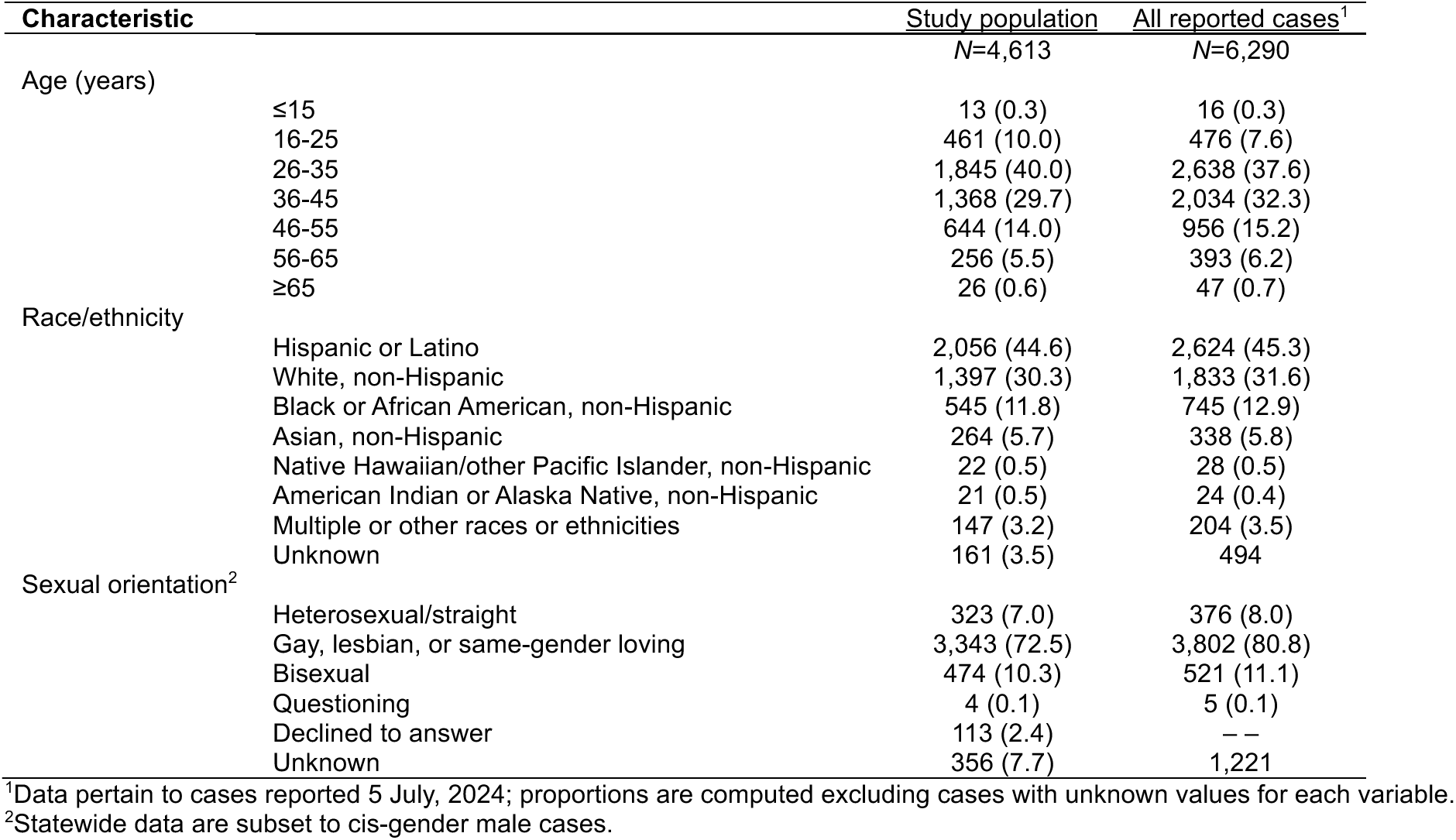
Demographic characteristics of the study population and mpox cases reported in California.

**Table S2:** Association of receipt of tecovirimat with progression to disseminated lesion occurrence.

| Vaccination status | Tecovirimat receipt | Pattern of lesion occurrence, *n* (%) |  | aOR (95% CI) |
| --- | --- | --- | --- | --- |
|  |  | Disseminated lesions across multiple anatomical regions | Lesions contained to a single anatomical region |  |
| Any vaccination status |  | *N*=3,045 | *N*=1,568 |  |
|  | Tecovirimat not received | 2,455 (80.6) | 1,326 (84.6) | ref. |
|  | Tecovirimat received | 590 (19.4) | 242 (15.4) | 1.08 (0.98, 1.20) |
| Unvaccinated |  | *N*=2,717 | *N*=1,131 |  |
|  | Tecovirimat not received | 2207 (81.2) | 955 (84.1) | ref. |
|  | Tecovirimat received | 510 (18.8) | 181 (15.9) | 1.06 (0.95, 1.18) |
| Any pre-exposure immunization |  | *N*=114 | *N*=286 |  |
|  | Tecovirimat not received | 89 (78.1) | 246 (86.0) | ref. |
|  | Tecovirimat received | 25 (21.9) | 40 (14.0) | 1.61 (0.95, 2.73) |
| Partial (1-dose) pre-exposure series |  | *N*=71 | *N*=169 |  |
|  | Tecovirimat not received | 56 (78.9) | 144 (85.2) | ref. |
|  | Tecovirimat received | 15 (21.1) | 25 (14.8) | 1.50 (0.77, 2.94) |
| Full (2-dose) pre-exposure series |  | *N*=43 | *N*=117 |  |
|  | Tecovirimat not received | 33 (76.7) | 102 (87.2) | ref. |
|  | Tecovirimat received | 10 (23.3) | 15 (12.8) | 1.77 (0.75, 4.17) |
| Immunization as post-exposure prophylaxis alone |  | *N*=214 | *N*=146 |  |
|  | Tecovirimat not received | 159 (74.3) | 125 (85.6) | ref. |
|  | Tecovirimat received | 55 (25.7) | 21 (14.4) | 1.15 (0.81, 1.64) |

CI: Confidence interval; aOR: adjusted odds ratio. We compute the aOR of participants' history of receiving tecovirimat comparing those who experienced comparing cases who experienced disseminated lesions to those who experienced lesions contained to a single region. We estimate the aOR via conditional logistic regression, matching participants on JYNNEOS vaccination history according to the categories defined in the left-most column.

**Table S3:** Association of JYNNEOS vaccination with number of anatomical regions where lesions occurred.

| Vaccination status | Adjusted odds ratio (95% CI) |  |  |
| --- | --- | --- | --- |
|  | All cases | HIV negative cases | Cases living with HIV |
| Unvaccinated | ref. | ref. | ref. |
| Any pre-exposure immunization | 0.43 (0.38, 0.48) | 0.36 (0.29, 0.44) | 0.50 (0.42, 0.59) |
| Partial (1-dose) pre-exposure series | 0.42 (0.36, 0.49) | 0.37 (0.30, 0.46) | 0.49 (0.39, 0.61) |
| Full (2-dose) pre-exposure series | 0.40 (0.33, 0.49) | 0.35 (0.27, 0.46) | 0.49 (0.39, 0.61) |
| Immunization as post-exposure prophylaxis alone | 0.79 (0.73, 0.85) | 0.75 (0.67, 0.84) | 0.84 (0.75, 0.93) |

CI: Confidence interval. We adjusted odds ratios addressing relative odds of vaccination for each additional anatomical region at which cases reported lesions being present, measured as a continuous parameter, via conditional logistic regression matching participants on HIV infection status.

**Table S4:**
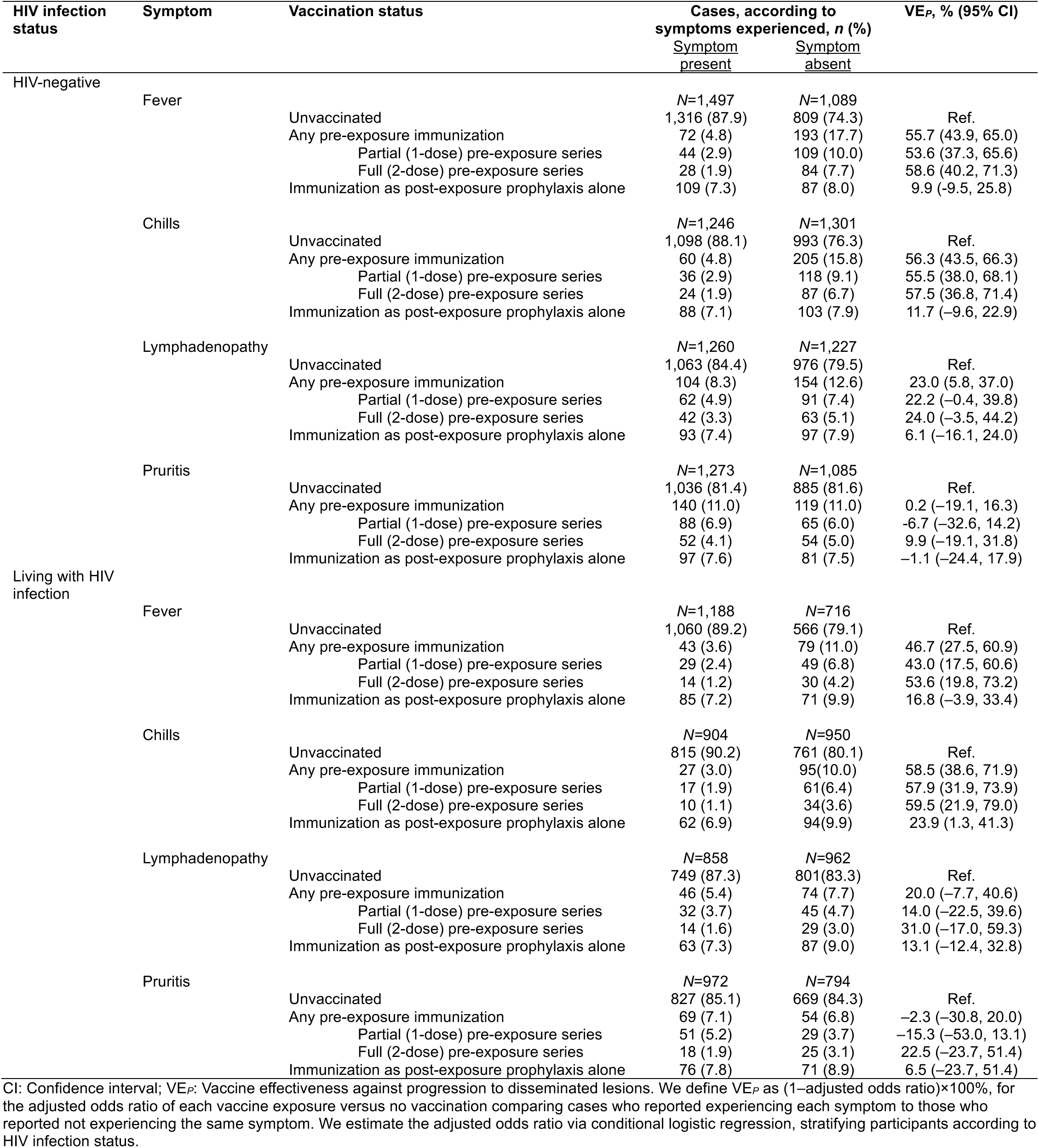
JYNNEOS vaccine effectiveness against progression (VE*_P_*) to illness involving fever, chills, lymphadenopathy, and pruritis, according to cases’ HIV infection status.

**Table S5:**
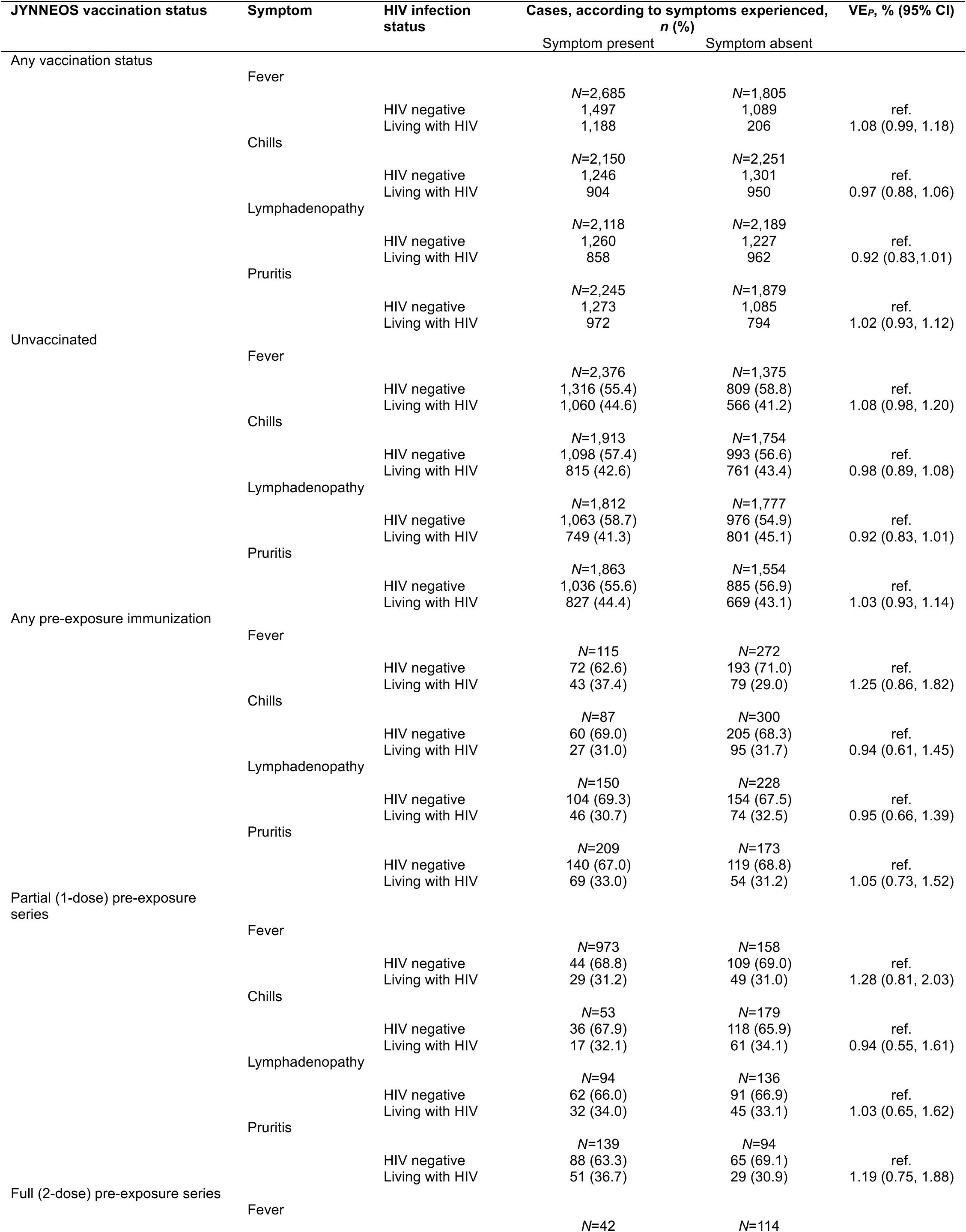
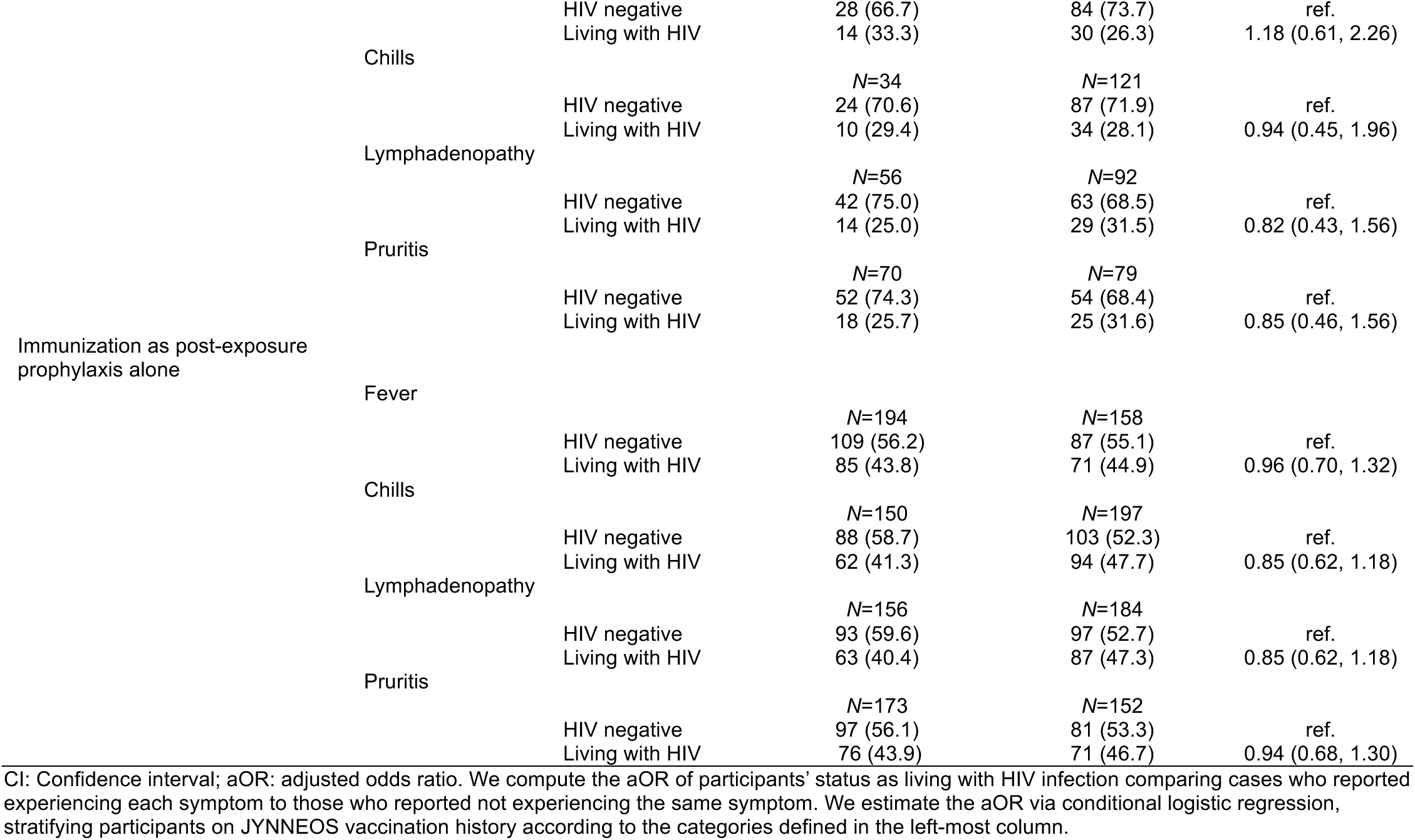
Association of HIV infection with progression to illness involving fever, chills, lymphadenopathy, and pruritis.

